# The development and validation of a survey to measure fecal-oral child exposure to zoonotic enteropathogens: The FECEZ Enteropathogens Index

**DOI:** 10.1101/2023.11.15.23298587

**Authors:** April M. Ballard, Regine Haardörfer, Betty Corozo Angulo, Matthew C. Freeman, Joseph N.S. Eisenberg, Gwenyth O. Lee, Karen Levy, Bethany A. Caruso

**Affiliations:** Department of Population Health Sciences, Georgia State University School of Public Health; Gangarosa Department of Environmental Health, Emory University Rollins School of Public Health; Department of Behavioral, Social, and Health Education Sciences, Emory University Rollins School of Public Health; Universidad Técnica Luis Vargas Torres de Esmeraldas; Department of Epidemiology, University of Michigan School of Public Health; Rutgers Global Health Institute and Department of Biostatistics and Epidemiology, Rutgers School of Public Health; Department of Environmental and Occupational Health Sciences, University of Washington School of Public Health; Hubert Department of Global Health, Emory University Rollins School of Public Health

## Abstract

Child exposure to animal feces and associated enteropathogens contribute to a significant burden of disease in low- and middle-income countries. However, there are no standardized, validated survey-based approaches to enable accurate assessment of child exposure to zoonotic enteropathogens. We developed and validated a survey-based measure of fecal-oral child exposure to zoonotic enteropathogens, the FECEZ Enteropathogens Index. First, we identified critical attributes of child exposure through in-depth interviews (IDIs) in Ecuador among individuals who care for animals (*n*=29) and mothers of children under two years old (*n*=58), and through a systematic review of existing exposure measures. Second, based on these findings, we developed a 105-question survey and administered it to 297 mothers with children under age five. Third, we refined the survey, using principal component analysis to determine the optimal number of components. The final index consisted of 34 items across two sub-domains: the child *Environment* and child *Behavior*. Lastly, we compared index scores to two commonly used, unvalidated measures of child exposure – maternal reported household animal ownership and presence of animal feces. Using the FECEZ Enteropathogens Index revealed varying degrees of exposure in our study population, with only two children having no exposure. In contrast, if we had used animal ownership or the presence of animal feces as a measure of exposure, 44% and 33% of children would have been classified as having no exposure, respectively. These common binary exposure measures may be inadequate because they do not provide sufficient information to identify the relative risk of zoonotic pathogen exposure. The FECEZ Enteropathogens Index overcomes this limitation, advancing our ability to assess exposure by quantifying the multiple components of child exposure to zoonotic enteropathogens with higher resolution. Additional testing and evaluation of the index is needed to ensure its reliability, validity, and cross-cultural equivalence in other contexts.

## Introduction

Enteric pathogens pose serious health risks for children under age five. In addition to acute diarrheal disease (the fifth leading cause of death among children under five years), persistent exposure and recurrent enteric infections are associated with environmental enteric dysfunction, and deficits in growth and cognitive development.[1–4] Enteric infections and their sequelae disproportionately affect children living in poverty in low- and middle-income countries (LMICs) due to inequitable access to healthcare, inadequate water and sanitation infrastructure, and widespread fecal contamination of the environment.[2, 5–15]

Exposure to animal feces is an important transmission route of enteropathogens,[16–20] particularly among children in LMICs where animals are ubiquitous and insufficient separation of animal feces from domestic spaces is well documented.[20–26] Many pathogens capable of infecting humans are transmissible via animal feces, some of which contribute significantly to the global burden of diarrheal disease. Although the specific attributable fraction of animal-sourced infections is unquantified, four pathogens that can be transmitted in animal feces (*Campylobacter* spp., *Cryptosporidium* spp., enteropathogenic *E. coli*, non-typhoidal *Salmonella*) are responsible for 28.3% of the estimated global diarrhea deaths in children under five years.[16] Global animal feces production greatly exceeds that of humans; livestock animal feces accounts for 80% of the global fecal load. A significant number of households have these animals onsite, where human contact with feces is high.[27]

Significant challenges remain in understanding the scope of child exposure to zoonotic enteropathogens, especially in the areas of highest risk. Such challenges are in part due to current approaches to exposure assessment, which are diverse and distal from exposure itself.[28, 29] Researchers assess exposure inconsistently with varied methods – including via survey, observation, and microbiology techniques – which limits comparisons across studies and settings. Existing measures also overwhelmingly assess a single attribute of exposure, typically related to animals (e.g., presence of animals) or the environment (e.g., presence of animal feces).[28, 29] Such approaches fail to account for the multiple factors that lead to exposure and do not capture more proximal factors (e.g., human contact with animals and animal feces) that play a central role in exposure. For example, the presence of animals or animal feces may not consistently be good proxies for exposure if children do not interact with the animals or their feces. Conversely, accounting only for child behavior does not capture whether animal-sourced contaminants are present. A standard measure that captures the multiple factors and conditions that contribute to exposure is therefore needed to improve the assessment of child exposure to zoonotic enteropathogens and to enable comparisons within and across communities.

In this study, we build upon existing measures of fecal-oral child exposure to zoonotic enteropathogens and address prevailing measurement limitations by developing and validating a survey-based measure, i.e., the FECEZ Enteropathogens Index. Here we report the sequential mixed methods approach we employed to develop the index, followed by the results from our measurement development and evaluation using data from northwestern coastal Ecuador.

## Methods

### Defining and conceptualizing child exposure to zoonotic enteropathogens

Following established practices for measurement development,[30–32] we offer a preliminary definition of our focal construct of interest, ‘child exposure to zoonotic enteropathogens’: ingestion of enteropathogens through direct and indirect contact with animals, animal feces, and fecal contamination. Alongside this definition, we developed a framework (Fig 1) adapted from the exposure science source-to-outcome continuum,[33] as well as our recent qualitative research and systematic review, that is the conceptual basis of the FECEZ Enteropathogens Index. The exposure science continuum – which includes source; contaminant; contaminated environmental media, objects, and surfaces; behavior, route, and outcome – delineates the specific elements that should be assessed to robustly measure exposure. Our framework considers exposure to be constituted by two distinct sub-domains, which are the critical attributes or characteristics of our focal construct. The *Environment* sub-domain includes the child’s household, compound, and interpersonal environment. This sub-domain focuses on sources of zoonotic enteropathogens (i.e., animals and their feces), the contaminant itself (i.e., enteropathogens in animal feces), and contaminated environmental media, objects, and surfaces. The *Behavior* sub-domain includes child behaviors inside and outside the household compound. This sub-domain focuses on interactions with potential sources, contaminants, and contaminated environmental media, objects, and surfaces that could lead to ingestion of zoonotic enteropathogens.

**Fig 1.**
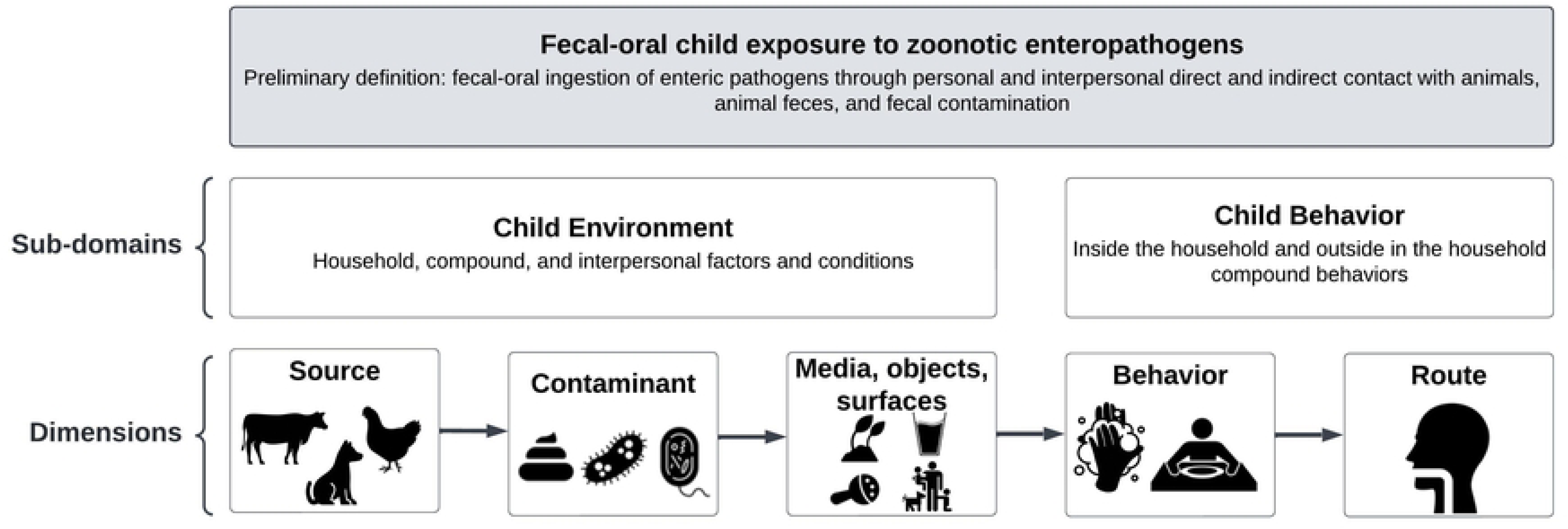
Conceptual framework of the focal construct of interest, fecal-oral child exposure to zoonotic enteropathogens.^a^. ^a^Construct: a well-defined and bounded subject of measurement; sub-domains: critical attributes or characteristics of the construct of interest; dimensions: characteristics or elements that constitute the construct of interest and its sub-domains [30, 31]

### Setting

Our study was conducted in multiple communities in northwestern coastal Ecuador as part of an ongoing birth cohort study, Enteropatógenos, Crecimiento, Microbioma, y Diarrea (referred to as ECoMiD).[34] The EcoMiD study assesses the impact of environmental exposures on enteric pathogen infections, gut microbiome composition, and development during the first two years of children’s lives. The region is primarily populated by Afro-Ecuadorians, mestizos, and some indigenous individuals. Data were collected in four ECoMiD sites: (1) Esmeraldas, the most urban site in the study area; (2) Borbón, a smaller town in Esmeraldas Province that serves as a commercial center; (3) rural villages near Borbón that are accessible by road; and (4) rural villages near Borbón that are accessible only by boat. Esmeraldas (population: 160,000 [35]) is a densely populated city and the capital of Esmeraldas Province. The city has the most access to water, sanitation, and infrastructure.[36] Borbón (population: 7,700 [35]), a town in the Esmeraldas Province, is located at the convergence of the Cayapas, Santiago, and Onzole rivers. The town has inadequate infrastructure for its size, including minimal water and sanitation infrastructure (e.g., untreated sewage, basic solid waste management).[36, 37] Approximately 125 small villages (population: 50-500 per village [35]) lie along the three rivers, some of which have access to Borbón via road (i.e., rural road communities) and have less developed infrastructure, though many have been recently connected to drinking water systems.[36, 37] Other villages are only accessible by river (i.e., rural river communities) and are comparatively more remote and lack centralized infrastructure.

### Overview of research design

We used a sequential mixed methods approach [38] to create and evaluate the index, following measure development best practices.[30–32] An overview of the three-phase approach is depicted in Fig 2 and subsequently detailed.

**Fig 2.**
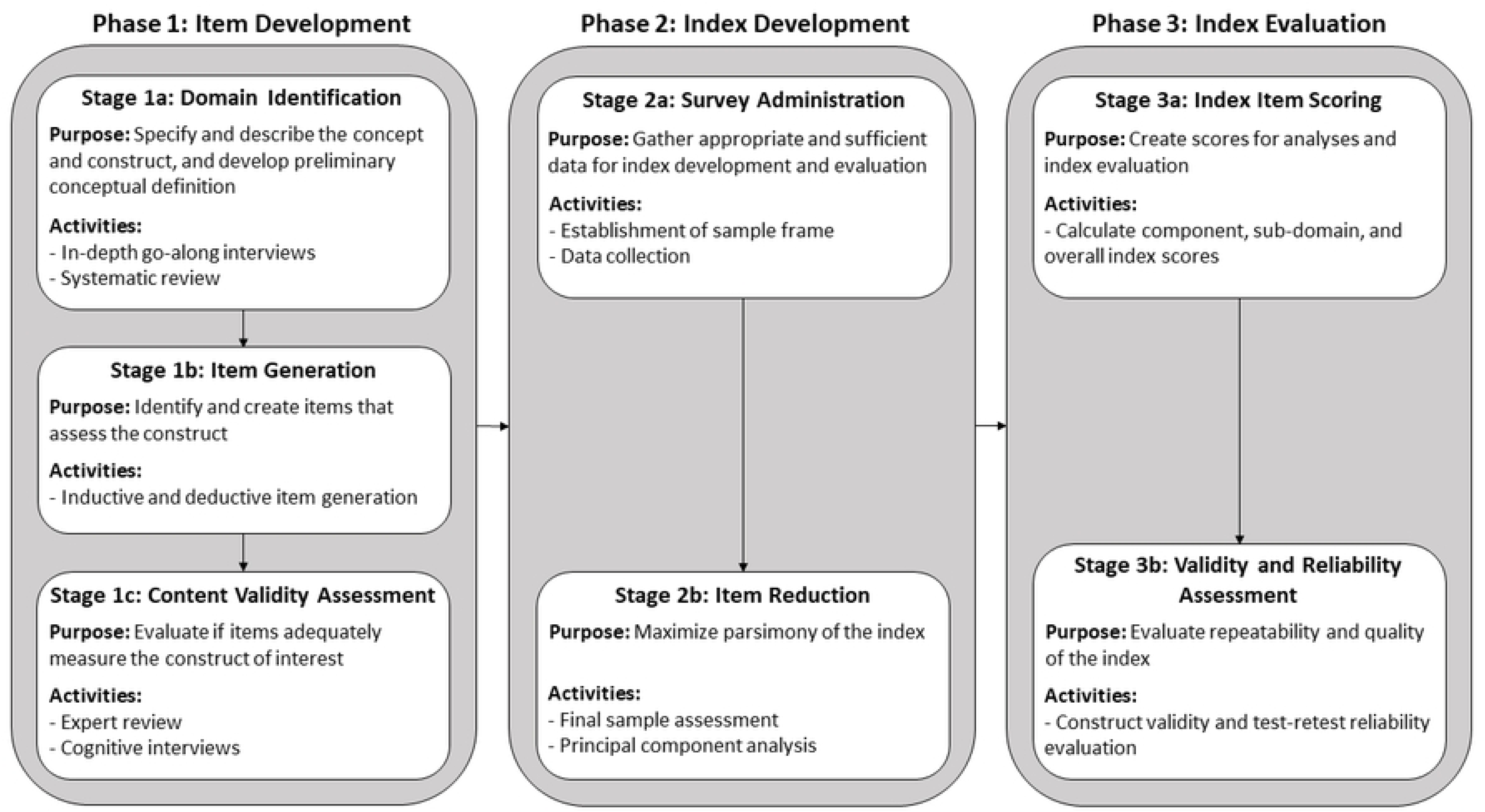
Schematic of the sequential mixed methods research design employed to create and evaluate the FECEZ Enteropathogens Index to measure child exposure to zoonotic enteropathogens.^a^. ^a^Items: survey questions that measure or reflect the construct of interest; content validity: the degree to which the items (i.e., the measurement tool) adequately reflect the construct of interest; construct validity: the degree to which the items accurately assess the construct of interest; test-retest reliability: the degree to which the items consistently measure the construct of interest over time [30, 31]

### Phase 1: Item Development

To identify content domains and generate potential index items (i.e., the survey questions that measure the construct of interest), we conducted in-depth interviews (IDIs) and relied on our recently published systematic review.[28] We finalized and evaluated the adequacy of the proposed items (i.e., content validity) through expert review and cognitive interviews with individuals similar to our survey target population, i.e., mothers of children 6-18 months of age in the study area.[30, 31]

#### Phase 1 Stage 1a: Domain Identification

##### In-depth go-along interviews

To define and conceptualize fecal-oral child exposure to zoonotic enteropathogens, we conducted 58 go-along, semi-structured IDIs with mothers of children 6-18 months of age enrolled in the on-going birth cohort study from January 21^st^ to April 21^st^, 2021. Go-along IDIs combine interviewing and participant observation, allowing the participant to actively engage with the spaces being discussed.[39, 40] We used purposive quota sampling to interview equal proportions of mothers who did (*n*=32) and did not (*n*=26) own animals to ensure a sufficient sample in each category to examine animal ownership’s impact on child exposure. Interviews queried conditions and maternal and child behaviors that could lead to child exposure to zoonotic pathogens, capturing details related to animals, environmental conditions, and behaviors on a typical day. To provide additional context about animals and environmental conditions, we conducted 29 interviews with individuals from the same communities who were not part of the cohort study and owned, cared for, and/or worked with various animals. Non-cohort IDIs queried how animals are cared for (e.g., feeding, animal feces management) and by whom, and decision-making about animal ownership and management. IDIs were conducted by co-author BCA, a qualitative researcher with more than 10 years of research experience who grew up and lives in the study area.

We analyzed IDI data to understand the scope of child exposure to zoonotic enteropathogens, reviewing transcript segments from relevant codes. We held debriefing meetings to conceptualize the constituents of exposure and then recorded the prominence or frequency that each constituent occurred in the sample to inform domain identification. Ultimately, we identified two content domains of exposure: the child’s environment and the child’s behaviors inside the home and outside the home in the household compound. Additional information about the qualitative methods, analyses, and findings are reported elsewhere.

##### Systematic review

To identify additional exposure attributes and characteristics and to make our index more generalizable to other settings, we drew upon our recently published systematic review in which we audit existing measures of human exposure to animal feces in LMICs. In our review, we identified 1,428 quantitative measures and categorized them based on what type of information they captured (e.g., presence of animals, animal fecal contamination of the environment, contact with animal feces) and where they fell along the exposure science continuum. These categorizations were then used alongside findings from the IDIs to solidify the content domains and dimensions and develop our conceptual framework. Detailed information about the methods and results of the systematic review are described elsewhere.[28, 29]

#### Phase 1 Stage 1b: Item Generation

Using our conceptual framework based on the exposure science continuum, IDIs, and the systematic review, we created an initial list of items for each sub-domain and dimension. We then reviewed existing measures captured via the systematic review to identify additional relevant items, focusing on comprehensiveness and alignment with our conception of child exposure.

#### Phase 1 Stage 1c: Content Validity Assessment

##### Expert review

To evaluate if proposed items adequately measure the construct (i.e., child exposure to zoonotic enteropathogens), we conducted three rounds of expert review. First, four co-authors (BAC, MCF, RH, KL) were sent draft items to assess the extent to which questions reflected the construct of interest and were appropriately worded, ordered, representative, and comprehensive. Second, we conducted a formalized expert review process with two other co-authors (JNSE, GOL) and two external experts who were asked to evaluate each item on a scale of 1-4 for representativeness and clarity and provide comments on representativeness, clarity, and comprehensiveness. Scores and comments for each item were used to identify items to be edited, deleted, or added. Third, we translated items from English to Spanish and the co-author (BCA) who conducted IDIs for this study reviewed and commented on items.

##### Cognitive interviews

As a final assessment of content validity, co-author BCA conducted 20 cognitive interviews with mothers in the study area who were not enrolled in the ECoMiD study from March 2^nd^-21^st^, 2022. The researcher asked participants to explain each question in their own words and how they determined their responses to questions to evaluate the suitability for the target population and potential sources of response error. Edits were made based on cognitive interview findings and debriefing meetings between co-authors AMB and BCA.

### Phase 2: Index Development

To collect data for index development and evaluation, we administered the revised items within a broader survey, and then used principal components analysis (PCA) to reduce the number of items to maximize parsimony.

#### Phase 2 Stage 2a: Survey Administration

##### Sampling frame

Although no consensus for optimal sample size exists for this cross sectional survey and subsequent analysis, 200-300 participants has previously been found to be adequate for performing PCA.[30, 41–43] Participant recruitment followed a random-walk method in the same neighborhoods where IDIs were conducted and where the ECoMiD study participants reside. An enumerator walked through neighborhoods and knocked on doors to screen participants for eligibility. To be eligible, the participant was required to be aged 18 or older, a mother to a child six months to five years old, and not a member of the ECoMiD cohort study. ECoMiD cohort participants were excluded to avoid research fatigue given their ongoing participation in various cohort activities. If an eligible individual consented to participate, the enumerator would administer the survey or make an appointment to return.

#### Data collection

The survey included the proposed measure items from Phase 1, as well as modules on mother and child demographics, maternal perception and knowledge about exposure to animals and animal feces, water and sanitation access, child health, and household characteristics. There were 52 proposed items designed to measure the two exposure sub-domains. Each item asked mothers how often a particular event or behavior occurred in the last week: never, rarely, sometimes, frequently (see S1 Table for full list of items). Items were ordered to build in intensity or proximity to exposure. For example, *Environment* items were asked first and items about child contact with animals and their feces were asked last. Items that assessed the presence of animals and animal feces in the child’s indoor and outdoor home environment included potential follow-up items based on responses, meaning mothers were asked a range of 52-105 questions (85 *Environment* items and 20 *Behavior* items). The survey was administered by a single enumerator using Open Data Kit (ODK) and an Android tablet. Prior to data collection, the enumerator completed one week of training by co-author AMB about the purpose of the survey, interview techniques, research ethics, and logistics. Survey data collection occurred from August 3^rd^ to September 30^th^, 2022. The total survey took an average of 30 minutes to complete. Participants received an assortment of household items, such as soap and toothpaste, as compensation for their time.

To facilitate the assessment of test-retest reliability (i.e., how consistent index scores are across time), we re-administered the survey sections with measure items to approximately 25% of participants within a three to seven day window.[30, 31] Repeat surveys took approximately 15 minutes to complete and were conducted among a random sample of those who agreed to a second visit. A 3-to-7-day window was selected because the items refer to ‘the past week’ and we wanted to capture responses within an overlapping period to reduce the likelihood of meaningful events or changes that would lead to different responses to survey items.

#### Phase 2 Stage 2b: Item Reduction

##### Final sample assessment

Prior to PCA, we removed items if 1) few respondents (<5%) reported a certain condition or behavior and 2) two items were highly correlated (*r* >0.90). In the latter case, one item was retained and the other was removed to reduce the number of initial items to a smaller subset of less-highly correlated items where logic and theory supported items interrelatedness.

To evaluate if each set of items were suitable for PCA, we conducted three statistical tests. First, Kaiser-Meyer-Olkin Measure of Sampling Adequacy [44] was used to determine the proportion of variance in variables that may be caused by underlying factors. We considered values above 0.50 overall and per item as adequate in demonstrating that PCA was useful to reduce the dimensionality of our data.[45, 46] Second, Bartlett’s Test of Sphericity [47] was used to test if the correlation matrix was an identity matrix, meaning variables are unrelated and therefore not suitable for PCA. We considered *p*-values less than 0.05 an indication that the correlation matrix was not an identity matrix.[46] Lastly, we calculated the determinant of the correlation matrix to assess if multicollinearity or singularity were issues in our data. We considered determinant values >0.00001 to indicate no issues.[46]

##### Principal component analysis

To determine the optimal number of components (i.e., linear combinations of the original variables that captures patterns of correlations related to an underlying factor and is derived through PCA [48]) that fit each sub-domain, we conducted categorical PCA with multivariate analysis with optimal scaling using the *princals* function from the *Gifi* package [49] in R Studio version 4.0.5 [50] on the raw ordinal data for the two subdomains separately. We used a linearly scaled fit (i.e., linear knots with no interior knots) to transform the ordinal values to be linearly scaled with equal distances between points, which aligns with the meaning behind the ordinal data (i.e., the number of days). To identify the number of principal components (PCs) to retain, we considered eigenvalues, a scree plot, and in part, theory based on our conceptualization of exposure. We used Kaiser’s Criteria (eigenvalues >1.0) and a scree plot to determine the ‘elbow’ point, which demarks where the eigenvalues go from exponential decay to a linear trend.[31, 48, 51] To decide on final solutions, we iteratively re-ran analyses to examine solutions with a varying number of components, balancing interpretability and the percent of variance explained to produce parsimonious, functional, and interpretable indices.[30, 31, 52] We also assessed item loadings and theoretical fit of each item within components. We decided *a priori* to conduct stepwise removal of items with loadings <0.40 and/or that were loaded on several components and did not theoretically make sense.[31, 48] After each item was removed, statistical tests and eigenvalues were re-assessed to make sure the remaining data were still suitable for PCA and that we were still assessing an appropriate number of components. The final component structures were assessed using knowledge of child exposure to zoonotic enteropathogens to ensure that items and components were appropriate and relevant.

### Phase 3: Index Evaluation

To create and evaluate the FECEZ Enteropathogens Index, we calculated scores based on PCA results and assessed scores construct validity and test-retest reliability in R studio version 4.0.5.[50]

#### Phase 3 Stage 3a: Index Item Scoring

To calculate PC scores, we used an unweighted approach and calculated the sum of responses for each final item using the original, ordinal values (i.e., 0 = never, 1 = rarely, 2 = sometimes, 3 = frequently).[30, 31] We calculated each components’ score by summing each item’s ordinal values together for items that loaded to that component in the final PCA solutions. If items were cross-loaded, the item was considered part of the component where the loadings were the largest.

To create index scores for substantive analysis, PC scores were used to calculate sub-domain (or sub-index) and overall index scores. We used summations of ordinal data to calculate index scores, as opposed to transformed scores from component loadings, to facilitate interpretability and index score comparisons in future research given that loadings will differ by study population. Higher scores indicate a greater frequency of occurrence.

#### Phase 3 Stage 3b: Construct Validity and Test-retest Reliability Assessment

To evaluate if the final index accurately assessed what it was designed to (i.e., construct validity), we conducted bivariate linear regression analyses to examine if the measure behaves as expected in relation to “known groups.”[30, 31] Specifically, we assessed whether scores for each component, sub-domain, and the overall index were significantly different by community type (i.e., urban, commercial center, rural road, rural river), hypothesizing that there would be detectable differences in exposure levels across the urban-rural gradient because the number and diversity of animals varies across the sites. We also investigated whether scores were significantly different by household animal ownership, as existing literature suggest that exposure may be higher among children in households with animals, and by child age, given that there are many developmental milestones that could impact exposure in the first five years of life.

To determine if the index provided a stable measure that can be used on repeat occasions (i.e., test-retest reliability),[30, 31] we calculated intraclass correlation coefficient (ICC) estimates and their 95% confidence intervals using the *ICC* function from the *psych* package [53] in R studio version 4.0.5[50] based on a single-rating, absolute agreement, two-way mixed effects model.[54] ICCs were calculated for each PC, the *Environment* and *Behavior* sub-indices, and the overall index for participants who were surveyed twice. We used the following guidelines to evaluate ICC values: <0.50 poor reliability, 0.50-0.75 moderate reliability, 0.76-0.90 good reliability, and >0.90 excellent reliability.[54]

### Ethics

Emory University (IRB # 00101202) and Universidad San Francisco de Quito Institutional (IRB # 2018-022M and 021-011M) Review Boards approved all study activities. Participants provided written consent prior to data collection.

## Results

### Participant Demographics

In total, we administered 297 surveys across the four study sites. Most households owned at least one type of animal (55.6%, *n*=165), with dogs and cats being the most common (Table 1). A quarter (*n*=75) of children were reported by mothers to have had a fever in the week prior, 13.5% (*n*=40) had diarrhea, and 7.7% (*n*=23) had vomited. Sex-disaggregated demographic characteristics are provided in S2 Table.

**Table 1.**
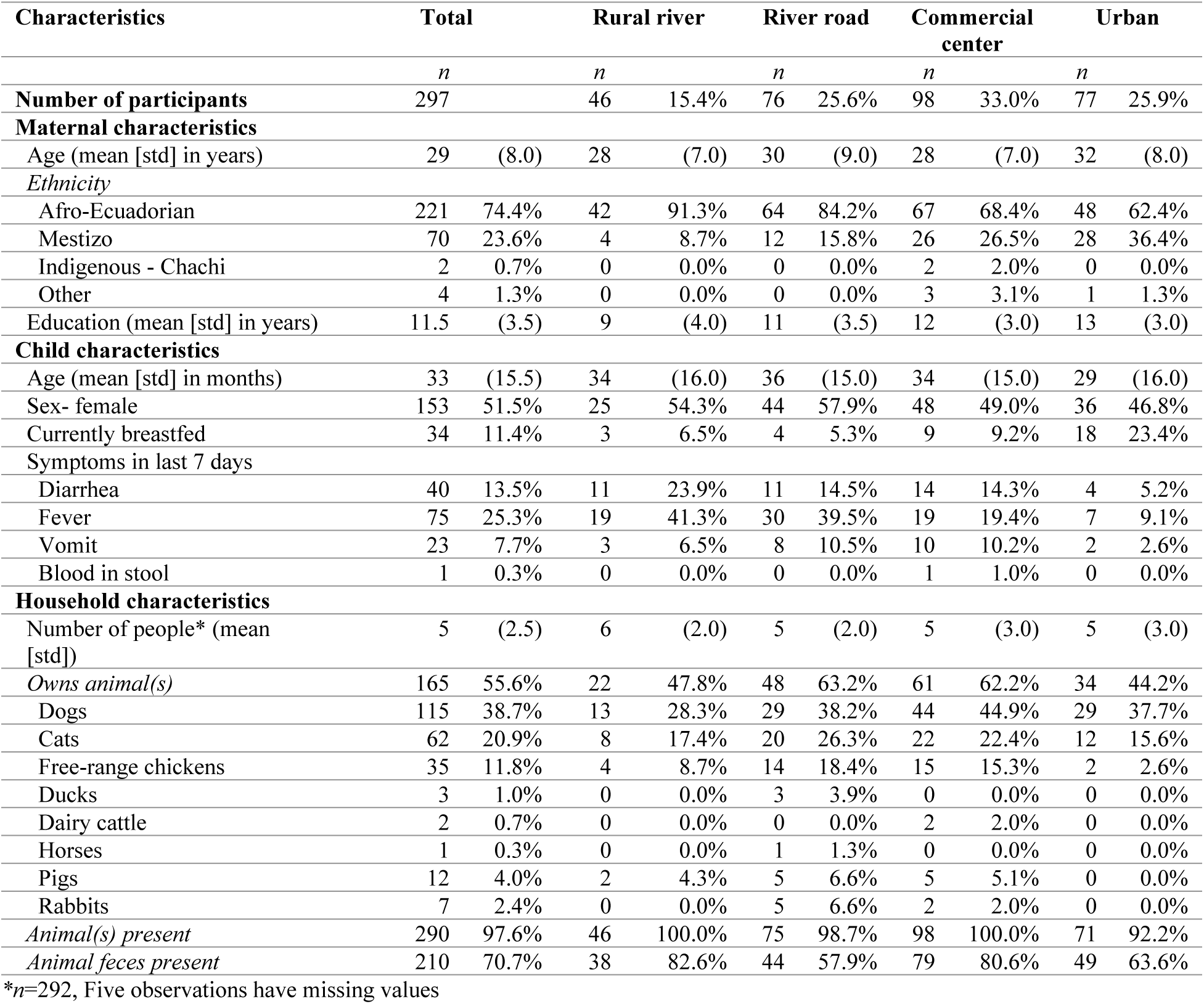
Maternal, child, and household characteristics for total sample and by the four study sites (*n*=297)

### Index Development

#### Final Sample Assessment

The survey included 105 items for potential inclusion in the final measures: 85 *Environment* items and 20 *Behavior* items. All participants responded “never” to 37 *Environment* items and one *Behavior* item, so they were eliminated due to their irrelevancy to this population. We eliminated 21 additional *Environment* items because they were near zero variance predictors, indicating they had limited relevance for the sample. Lastly, two *Behavior* items were eliminated because they were highly correlated and theoretically similar to two retained items. Specifically, items that captured children playing in sand outside the house and children putting sand in their mouths were removed because they were highly correlated with children playing in dirt or soil outside the house and children putting dirt or soil in their mouths (*r*>0.90). PCA was therefore conducted with 27 *Environment* items and 17 *Behavior* items to create two sub-indices. Distributions of item responses are in S3 Table. Items that were omitted and reasons for omission are in S1 Table.

#### Principal Component Analysis

Our three pre-analysis tests indicated that remaining data for each sub-domain were suitable for PCA. For the *Environment* and *Behavior* items, the Kaiser-Meyer-Olkin measure values were 0.71 and 0.75, respectively, indicating acceptable sampling adequacy. The Bartlett’s Test of Sphericity revealed that the between-item correlations were sufficient for PCA for both sets of items (*Environment* items: K-squared = 2094.8, degrees of freedom (df) = 19, *p*-value <0.001; *Behavior* items: K-square = 2079.4, df = 13, *p*-value <0.001). There were also no issues with multicollinearity or singularity; determinants of the correlation matrices were 0.002 and 0.01 for the *Environment* and *Behavior* items, respectively.

#### Child Environment sub-domain

For the *Environment* sub-domain, we determined that a five-component solution best suited the data theoretically, based on a screeplot, Kaiser’s Rule (eigenvalues >1.0), and the amount of variance explained by PCs. Seven *Environment* items were omitted during analyses due to loadings <0.40 and/or cross-loading that was not interpretable (S1 Table). The final *Environment* sub-index included 20 items and explained 57% of the variance (Table 2). The fit appeared to be good based on the interpretability of the PCs, a loss value (i.e., a measure of goodness of fit) of 0.89, and a solution being obtained with 43 iterations. Of note, lower loss values and fewer iterations indicate better fits, though no exact thresholds exist.

**Table 2.** Eigen values, explained variance, cumulative explained variance, and component loadings for child *Environment* sub-domain PCA solution.

| **Solution characteristics** | **PC1** | **PC2** | **PC3** | **PC4** | **PC5** |
| --- | --- | --- | --- | --- | --- |
| Eigen value | 3.76 | 2.35 | 2.21 | 1.73 | 1.26 |
| Variance explained by PC | 0.19 | 0.12 | 0.11 | 0.09 | 0.06 |
| Cumulative variance explained | 0.19 | 0.31 | 0.42 | 0.51 | 0.57 |
| **Solution items** |  |  |  |  |  |
| Mother personally feeds or gives water to an animal | **-0.79** | -0.08 | -0.13 | -0.02 | -0.03 |
| Mother personally cleans the habitat or place where an animal sleeps and/or defecates | **-0.75** | 0.09 | -0.26 | 0.02 | -0.19 |
| Mother personally bathes, cleans, or grooms an animal | **-0.71** | 0.12 | -0.33 | -0.00 | -0.05 |
| Mother personally touches or play with an animal | **-0.70** | 0.07 | -0.30 | 0.00 | 0.03 |
| Mother personally eliminates or cleans the poop of an animal | **-0.70** | -0.12 | -0.29 | 0.15 | -0.24 |
| Dogs enter the house | **-0.53** | 0.30 | -0.03 | -0.28 | 0.32 |
| Free-range chickens spend time outside near the house | -0.27 | **-0.77** | 0.12 | -0.06 | 0.00 |
| Free-range chickens enter the house | -0.12 | **-0.72** | 0.05 | -0.05 | -0.01 |
| Free-range chicken poop outside the house near or in the yard | -0.24 | **-0.70** | 0.20 | 0.11 | 0.03 |
| Free-range chicken poop inside the house | -0.10 | **-0.56** | 0.11 | 0.15 | 0.02 |
| Cats enter the house | -0.37 | 0.22 | **0.76** | 0.23 | 0.02 |
| Cats spend time outside near the house | -0.25 | 0.22 | **0.64** | 0.29 | -0.13 |
| Cats sleep inside the house | -0.40 | 0.21 | **0.60** | 0.16 | 0.11 |
| Cat poop outside the house near or in the yard | -0.15 | -0.05 | **0.40** | 0.30 | -0.26 |
| Dairy cattle spend time outside near the house | -0.10 | -0.11 | 0.26 | **-0.70** | -0.05 |
| Dairy cattle poop outside the house near or in the yard | -0.00 | -0.02 | 0.25 | **-0.70** | -0.08 |
| Household member apart from mother and child under 5 years works or cares for an animal | -0.16 | -0.08 | 0.37 | **-0.52** | 0.00 |
| Dogs sleep inside the house | -0.44 | 0.23 | 0.06 | -0.12 | **0.58** |
| Dog poop outside the house near or in the yard | -0.17 | 0.24 | 0.07 | -0.22 | **-0.57** |
| Dogs spend time outside near the house | 0.04 | 0.17 | -0.01 | -0.13 | **-0.53** |

PC = Principal component; bold numeric values indicate item loading to the specific PC

The *Environment* sub-domain PCA yielded strong loadings onto five interpretable components, each listed with the proportion of variance accounted for: maternal factors (19%), free-range chicken factors (12%), cat factors (11%), dairy cattle factors (9%), and dog factors (6%) (Table 2). The PCs broadly corresponded to our two initially hypothesized *Environment*-related dimensions: the child’s household and compound environment and the child’s interpersonal environment. Specifically, PCs 2-5 included items related to specific species of animals and their feces. The item about other household members working with or caring for animals loaded on PC4 with dairy cattle items, which could represent farming communities/households where dairy cattle are present and family’s own, work with, and/or care for them. PC1 included items about maternal behaviors and interactions with animals and their feces. The items about dogs entering and dogs and cats sleeping in the household also loaded on this component, which likely indicates that mothers interact with dogs and cats and their feces that enter or sleep inside their house. Dogs entering the household loaded to PC1 (maternal factors) and not to PC5 (dog factors), which could be indicative of dogs specifically entering houses to be fed, bathed, groomed, or played with and contributing to interpersonal environmental contamination.

#### Child Behavior sub-domain

For the *Behavior* sub-domain, a two-component solution best suited the data. Three additional items were omitted during analyses due to loadings <0.40 and/or cross-loading (S1 Table). The final *Behavior* sub-index included 14 items, explaining 42% of the variance (Table 3). The fit appeared adequate with a loss value of 0.79 and a solution obtained with 12 iterations, and based on the PCs interpretability.

**Table 3.** Eigen values, explained variance, cumulative explained variance, and component loadings for child *Behavior* sub-domain PCA solution.

| **Solution characteristics** | **PC1** | **PC2** |
| --- | --- | --- |
| Eigen value | 3.34 | 2.52 |
| Variance explained by PC | 0.24 | 0.18 |
| Cumulative variance explained | 0.24 | 0.42 |
| **Solution items** |  |  |
| Child puts objects or toys that had contact with the dirt outside your house in their mouth | **0.73** | -0.25 |
| Child plays outside the house without shoes on | **0.67** | 0.13 |
| Child puts dirt or soil in their mouth | **0.59** | -0.28 |
| Child plays in dirt or soil outside the house | **0.59** | 0.05 |
| Child puts objects or toys that had contact with the floor inside your house in their mouth | **0.58** | -0.20 |
| Child plays outside the house in an area where an animal lives or sleeps | **0.56** | 0.12 |
| Child puts shoes in their mouth | **0.53** | -0.32 |
| Child plays with or carries around shoes like a toy | **0.52** | 0.02 |
| Child touches or plays with an animal | **0.45** | 0.34 |
| Child cleans or helps others clean the habitat or place where an animal sleeps and/or defecates | 0.16 | **0.76** |
| Child bathes, cleans, or grooms or helps others bathe, clean, or groom an animal | 0.22 | **0.75** |
| Child feeds or gives water or helps others feed or give water to an animal | 0.29 | **0.67** |
| Child cares for or helps others care for an animal that was sick | 0.02 | **0.56** |
| Child touches, removes, or cleans animal poop | 0.11 | **0.54** |

PC = Principal component; bold numeric values indicate item loading to the specific PC

The *Behavior* sub-domain PCA yielded strong loadings onto two interpretable components, each listed with the proportion of variance accounted for: play and mouthing behaviors (24%), and animal caregiving and feces management behaviors (18%) (Table 3). The components broadly corresponded to our structuring of questions that build in proximity to exposure to animals and their feces. Specifically, PC1 is comparatively more distal from exposure, including items about child play in potentially risky environments and mouthing potentially contaminated objects. PC2 includes items of increasing proximity to exposure, specifically caring for or helping others care for animals and interacting with animal feces.

### Index Evaluation

#### Index Item Scoring

The mean overall FECEZ Enteropathogens Index score was 27.21 (standard deviation [SD]: 12.75) out of 102 (Table 4). The average *Environment* sub-domain and *Behavior* sub-domain scores were 14.63 (SD: 8.57) out of 60 and 12.57 (SD: 7.10) out of 42, respectively. *Environment* and *Behavior* sub-domain scores were moderately positively associated, *r*(295) = 0.31, *p* <0.01 (S2 Fig). Histograms for sub-domain and overall index scores are provided in S1-S3 Figs. On average, scores were highest among children living in the rural river study site (Table 5). Children in the urban study site had the lowest average scores except for *Environment* PC5 (dog factors), signifying less child interaction with animals and their environment and less presence of animals and their feces, apart from dogs.

**Table 4.** Means and standard deviations of scores for Principal Components (PCs), Child *Environment* and *Behavior* sub-domains, and the overall FECEZ Enteropathogens Index (n=297)

|  | No. of items | Possible Score range | Mean score (std) |
| --- | --- | --- | --- |
| **Total** | 34 | 0-102 | 27.21 (12.75) |
| *Child Environment sub-total* | 20 | 0-60 | 14.63 (8.57) |
| PC1: Maternal factors | 6 | 0-18 | 5.42 (5.64) |
| PC2: Free-range chicken factors | 4 | 0-12 | 1.48 (2.67) |
| PC3: Cat factors | 4 | 0-12 | 2.92 (3.35) |
| PC4: Dairy cattle factors | 3 | 0-9 | 0.30 (0.96) |
| PC5: Dog factors | 3 | 0-9 | 4.52 (1.98) |
| *Child Behavior sub-total* | 14 | 0-42 | 12.57 (7.10) |
| PC1: Play and mouthing behaviors | 9 | 0-27 | 11.99 (6.55) |
| PC2: Animal caregiving and feces behaviors | 5 | 0-15 | 0.58 (1.72) |

**Table 5.** Component, sub-domain, and overall FECEZ Enteropathogens Index scores by community type and animal ownership, and child age.

|  | Mean score (sd) by community type |  |  |  | Mean score (sd) by animal ownership |  |  | Mean score (sd) by child age in months (mths) |  |  |  |
| --- | --- | --- | --- | --- | --- | --- | --- | --- | --- | --- | --- |
|  | Urban<br>(n=77, ref.) | Commercial center<br>(n=98) | Rural road<br>(n=76) | Rural river<br>(n=46) | None<br>(n=132, ref.) | 1 type<br>(n=110) | >1 type<br>(n=55) | 6-12 mths<br>(n=39, ref.) | 13-18 mths<br>(n=22) | 19-24 mths<br>(n=43) | 25-60 mths<br>(n=193) |
| **Total** | 18.10<br>(11.55) | **29.07**<br>**(11.45)** | **29.82**<br>**(11.12)** | **34.15**<br>**(12.05)** | 20.52<br>(11.16) | **30.05**<br>**(10.63)** | **37.55**<br>**(11.25)** | 19.64<br>(11.98) | **30.50**<br>**(12.63)** | **27.79**<br>**(13.02)** | **28.23**<br>**(12.40)** |
| *Child Environment sub-domain* | 10.32<br>(8.19) | **15.82**<br>**(8.66)** | **16.28**<br>**(8.13)** | **16.61**<br>**(7.40)** | 9.04<br>(6.06) | **16.91**<br>**(6.69)** | **23.51**<br>**(7.50)** | 11.87<br>(8.44) | 14.82<br>(7.90) | 15.35<br>(9.22) | **15.01**<br>**(8.48)** |
| PC1: Maternal factors | 4.14<br>(5.85) | 4.98<br>(5.02) | **6.86**<br>**(5.67)** | **6.15**<br>**(6.03)** | 2.02<br>(3.23) | **7.54**<br>**(5.52)** | **9.36**<br>**(5.87)** | 3.97<br>(5.35) | 5.64<br>(6.12) | 5.12<br>(5.56) | 5.76<br>(5.64) |
| PC2: Free-range chicken factors | 0.16<br>(0.96) | **1.39**<br>**(2.32)** | **2.03**<br>**(3.27)** | **2.96**<br>**(3.19)** | 0.85<br>(2.24) | **1.63**<br>**(2.63)** | **2.67**<br>**(3.23)** | 1.18<br>(2.55) | 1.95<br>(2.84) | 1.49<br>(2.59) | 1.48<br>(2.71) |
| PC3: Cat factors | 1.64<br>(2.79) | **3.54**<br>**(3.22)** | **3.43**<br>**(3.82)** | **2.87**<br>**(3.18)** | 2.04<br>(2.56) | 2.62<br>(3.35) | **5.62**<br>**(3.70)** | 1.95<br>(2.86) | 2.05<br>(2.50) | **4.02**<br>**(3.61)** | 2.96<br>(3.41) |
| PC4: Dairy cattle factors | 0.00<br>(0.00) | **0.89**<br>**(1.49)** | 0.04<br>(0.34) | 0.00<br>(0.00) | 0.09<br>(0.47) | 0.25<br>(0.84) | **0.91**<br>**(1.62)** | 0.05<br>(0.32) | **0.55**<br>**(1.14)** | 0.23<br>(0.72) | 0.34<br>(1.06) |
| PC5: Dog factors | 4.39<br>(2.23) | **5.02**<br>**(1.51)** | 3.92<br>(1.94) | 4.63<br>(2.25) | 4.04<br>(1.97) | **4.87**<br>**(1.92)** | **4.95**<br>**(1.91)** | 4.72<br>(2.18) | 4.64<br>(1.89) | 4.49<br>(2.15) | 4.47<br>(1.92) |
| *Child Behavior sub-domain* | 7.78<br>(5.19) | **13.26**<br>**(6.07)** | **13.54**<br>**(7.19)** | **17.52**<br>**(7.28)** | 11.48<br>(7.10) | 13.15<br>(6.89) | **14.04**<br>**(7.22)** | 7.77<br>(7.58) | **15.68**<br>**(7.82)** | **12.44**<br>**(6.56)** | **13.22**<br>**(6.63)** |
| PC1: Play and mouthing behaviors | 7.65<br>(4.92) | **12.36**<br>**(5.55)** | **12.92**<br>**(6.42)** | **16.96**<br>**(6.88)** | 11.35<br>(6.96) | 12.40<br>(6.22) | 12.73<br>(6.17) | 7.67<br>(7.46) | **15.18**<br>**(6.89)** | **12.23**<br>**(6.41)** | **12.45**<br>**(5.99)** |
| PC2: Animal caregiving and feces behaviors | 0.13<br>(0.77) | **0.90**<br>**(2.01)** | 0.62<br>(1.84) | 0.59<br>(1.86) | 0.14<br>(0.71) | **0.75**<br>**(1.68)** | **1.31**<br>**(2.85)** | 0.100<br>(0.50) | 0.50<br>(1.92) | 0.21<br>(0.71) | **0.77**<br>**(1.97)** |

Bold numeric values indicate $p$ -value $\leq 0.05$ for bivariate linear regression between scores and community type, animal ownership, and child age

#### Validity and Reliability Assessment

The PC, sub-domain, and total index scores differed significantly by community type (Table 5), indicating good construct validity (i.e., known-groups validity). As hypothesized, there was a statistically significant difference between scores across study sites. Children in rural river and rural road communities and the commercial center had significantly higher *Environment* sub-domain, *Behavior* sub-domain, and total index exposure scores compared to urban-residing children. There were also significant differences across PCs, though these differences varied by component as expected. For example, children in rural river and rural road communities and the commercial center had significantly higher scores related to free-range chickens (i.e., *Environment* PC2) compared to urban-residing children, which was expected given the sparseness of free-range chickens in the urban study site. However, only children in the commercial center study site had significantly higher scores related to dogs (i.e., *Environment* PC5), indicative of the presence of dogs and their feces across all study sites.

Children residing in households that owned at least one type of animal had significantly higher *Environment* sub-domain, *Behavior* sub-domain, and total index exposure scores compared to children in households that did not own any animals, as hypothesized. There were significant differences across PCs, though differences varied by component, as expected. For example, children in households that owned animals had significantly higher scores related to maternal factors (i.e., *Environment* PC1) compared to those residing in households without animals, indicative of the absence of animals in the household for mothers to interact with and care for. Conversely, there was not a significant difference between child play and mouthing scores (i.e., *Behavior* PC1), which was expected given that child play in potentially risky environments and mouthing potentially contaminated objects is not dependent on household animal ownership.

There were minimal differences in *Environment* sub-domain scores by child age, and there were significant differences between the *Behavior* sub-domain and *Behavior*-related PCs, as expected. For example, children who were 25-60 months of age had significantly higher scores related to animal caregiving and feces behavior (i.e., *Behavior* PC2) compared to children 6-12 months of age, which is expected given child developmental phases. There were also significant differences in overall exposure scores by child age, as hypothesized. Children become more mobile and active as they age, increasing events that could contribute to exposure.

ICC values, based on data from participants who were surveyed twice (*n*=66), for sub-domain and the overall index indicated good test-retest reliability (Table 6). The *Environment* sub-index, *Behavior* sub-index, and overall index had ICC values of 0.79, 0.81, and 0.81, respectively. ICC values for PCs showed moderate to excellent reliability. All five *Environment* sub-domain PCs had moderate to excellent test-retest reliability. For the *Behavior* sub-index, PC1 had good reliability (ICC: 0.86) and PC2 had moderate reliability (ICC: 0.66).

**Table 6.** Component, sub-domain, and overall FECEZ Enteropathogens Index scores intraclass correlation coefficient [ICC] estimates.

|  | ICC estimates |
| --- | --- |
|  | Kappa (95% CI) |
| **Total** | 0.81 (0.70, 0.88) |
| *Child Environment sub-domain* | 0.79 (0.68, 0.86) |
| PC1: Maternal factors | 0.87 (0.80, 0.92) |
| PC2: Free-range chicken factors | 0.75 (0.63, 0.84) |
| PC3: Cat factors | 0.83 (0.73, 0.89) |
| PC4: Dairy cattle factors | 0.51 (0.31, 0.67) |
| PC5: Dog factors | 0.70 (0.54, 0.81) |
| *Child Behavior sub-domain* | 0.81 (0.70, 0.88) |
| PC1: Play and mouthing behaviors | 0.86 (0.79, 0.92) |
| PC2: Animal caregiving and feces behaviors | 0.66 (0.49, 0.78) |

## Discussion

The FECEZ Enteropathogens Index – a valid and reliable two-domain, 34-item survey-based measure – advances our ability to assess exposure by quantifying the multiple components of child exposure to zoonotic enteropathogens with higher resolution. The measure produces a composite, continuous value that allows for the assessment of relative intensity of and variation in exposure and may decrease the likelihood of exposure misclassification. This tool is an initial step towards developing a universal index to compare exposure across populations. Below we describe the major strengths of the FECEZ Enteropathogens Index, including its integration of multiple factors to assess exposure and its ability to quantify the degree of exposure. We then outline the implications for research and practice, offering recommendations for the application of our tool and for future research to further refine the measure to ensure applicability to a broader context.

The FECEZ Enteropathogens Index revealed that exposure to zoonotic enteropathogens was ubiquitous among children in northwestern coastal Ecuador, a finding that would have been masked with the use of commonly used measures. Specifically, only two children were not exposed (i.e., had a score of zero). In contrast, if we had used animal ownership or the presence of animal feces as a measure of exposure in this study, 44% (*n*=132) and 33% (*n*=87) of children would have been classified as having no exposure, respectively. The assessment of child exposure using the presence of animals would have been more closely aligned the FECEZ index measure, but merely noting the presence of animals provides substantially less information compared to our index, which captures many factors to comprehensively assess exposure. Importantly, our index includes an entire domain with items to assess child behavior – a novel feature given that only 9% of existing exposure measures identified by our systematic review incorporated human behavior [28] – as well as multiple environmental factors, which are potential sources of enteropathogens (e.g., presence of specific animals and their feces) and pre-requisites of exposure.

The FECEZ Enteropathogens Index also captured exposure heterogeneity among our study population, a level of detail that would have been missed with typical binary measures of exposure. We found significant differences in the degree of exposure across communities and by household animal ownership and age. Commonly used binary measurements of zoonotic exposure [28] do not capture this variation and provide less information than a multidimensional measure like our FECEZ Enteropathogens Index. For example, binary measures such as the presence of animal feces or household animal ownership are unable to identify risk gradients among children and have the potential to introduce bias by misclassifying some children as unexposed when they in fact have some level of exposure.[55]

### Implications for research and practices

Child exposure scores produced using the FECEZ Enteropathogens Index were reliable and valid, suggesting that the measure functions well and can be tested more broadly. We found that the domain-specific and overall indices had good test-retest reliability, and exposure scores were significantly different across known-groups, demonstrating construct validity. Expert review and cognitive interviews during the item development phase strengthened the index’s ability to adequately measure exposure (i.e., content validity). Engaging urban and rural communities with different types of animals and husbandry practices increases the final measure’s generalizability, as does our use of existing exposure measures to create and evaluate the index.

Researchers and practitioners across sectors can use the 34-item index that we developed in this study to assess child exposure in several ways. For example, the index can inform program design by assessing baseline levels of exposure by sub-domain to identify areas for targeted intervention. The sub-domains can also be combined to comprehensively measure child exposure and identify inequities and vulnerable sub-populations within communities. Throughout program implementation, measurement using this index can enable the monitoring of exposure trends over time with higher resolution, supplying critical data for the development and evaluation of interventions. Such data can, in turn, facilitate the identification of upstream determinants of exposure and the assessment of the consequences of exposure among children.

Prior to implementing the FECEZ Enteropathogens Index in new locations, we offer two specific recommendations for researchers and practitioners. These steps are necessary for the measure’s use until its reliability and validity have been demonstrated across diverse contexts. However, researchers and practitioners may use the current version of the measure if the context is similar to northwestern coastal Ecuador, or use the index as a starting point for other local contexts.[56] First, we suggest evaluating content validity through cognitive interviews. During interviews, participants should be queried about the presence of feces from the types of animals that are relevant to the research context and known to be of high concern for pathogens transmitted in animal feces.[16] This step is crucial for ensuring a comprehensive assessment of exposure across settings because the FECEZ Enteropathogens Index consists of causal or formative indicators (as indices do generally [31, 57, 58]), meaning the absence of specific types of animals in this Ecuadorian study does not mean that they are not part of the construct, ‘child exposure to zoonotic enteropathogens.’ For example, we found that dogs, dairy cattle, cats, and free-range chickens were contributing to child exposure in Ecuador, which are all known to transmit four of the five pathogens that have been identified as of highest concern for pathogens transmitted in animal feces (i.e., *Campylobacter* spp., non-typhoidal *Salmonella*, *Cryptosporidium*, and *Toxoplasma gondii*).[16] However, other animals can also transmit these pathogens (e.g., swine can transmit non-typhoidal *Salmonella* and *Cryptosporidium*), and many other pathogens may have significant transmission potential through various types of animal feces (e.g., shiga toxin *E. coli*, *Toxocara canis*/*Toxocara cati*).[16] A list of animals to consider based on our research and existing literature is publicly available on OSF.[59]

Second, when applying the index in new locations, we recommend that researchers and practitioners conduct a PCA to test the component structure and assess test-retest reliability and content and construct validity. Such testing will explore the index’s external validity and help with progress toward the development of a global index. Survey items, scoring instructions, and detailed recommendations for the assessment of reliability and validity are publicly available on OSF.[59]

### Strengths and Limitations

A strength of this research is its’ multi-phase, rigorous approach to measurement development and evaluation. Use of qualitative interviews, a systematic review, expert review, and cognitive interviews for item development strengthen the measure’s content validity. Items serve as proxies to capture the components of exposure and many exposure pathways, improving upon existing exposure measures that only assess one aspect of exposure. However, this measure is limited by its inability to be a proxy for every potential animal feces exposure pathway. The measure does not directly assess pathways related to food, flies, and fluids (water), which are challenging to assess through survey and were therefore not included. Still, assessment of the final measure demonstrates good construct validity and test-retest reliability. Additionally, we were able to collect data from four study sites that differ, for example, by size, infrastructure, and livelihood systems. The diverse sample strengthens the external validity of the final measure, although testing in different populations in varied geographic locations is needed to further assess the measure’s generalizability. A limitation is our inability to examine how the measure performs compared to existing measures (i.e., concurrent validity), which we were unable to do because there is not a “gold standard” measure of child exposure to zoonotic enteropathogens. Lastly, our unweighted scoring approach may be less precise than a weighted approach that allows item scores to be based on their contribution to the component or factor. However, studies have found that weighting only improves precision moderately, makes interpretation challenging, and inhibits across-study comparisons.[31, 48] Producing unweighted scores is more user friendly, and will facilitate the standardization of exposure measurement and comparison of scores across studies.

## Conclusion

The FECEZ Enteropathogens Index advances our ability to assess exposure by quantifying the multiple components of child exposure to zoonotic enteropathogens with higher resolution. Researchers and practitioners may use the current version of the measure if the context is similar to the current study. Additional rigorous testing and evaluation of the index in different settings would ensure its reliability, validity, and cross-cultural equivalence in other contexts. Cognitive interviews and field testing of the tool should be conducted in diverse geographic locations as part of the ongoing iterative process aimed at establishing a global, standard exposure measure. The FECEZ Enteropathogens Index will be suitable for widespread use once its performance has been evaluated across multiple settings.[56] The advent of such a measure could impact on our understanding of child zoonotic enteropathogen exposures and can offer valuable evidence for the development of community’s public health and policy agendas.

## Data Availability

All relevant data are within the paper, its Supporting Information files, or can be found on OSF at the following URLs: https://osf.io/jkh3u/ and https://osf.io/d2nxc/. For more information, contact April Ballard.

https://osf.io/jkh3u/

## Acknowledgements

We would like to thank local field staff, especially in Borbón, for their help with recruiting, logistics, and data collection for this project; and Nick Laramee and Jayden Pace Gallagher for their contributions to the qualitative and systematic review phases of this study; and all the participants in Ecuador that made this project possible. We also thank Drs. Frederica Lamar and Sarah McKune for serving as external reviewers for the formalized expert review process.

## Supporting information

**Table S1. Table of survey items grouped by sub-domain and post-PCA results.**

**Table S2. Child sex-disaggregated demographic characteristics (n=297).**

**Table S3. Child exposure item frequencies (n=297).**

**Fig S1. Correlation plot between child behavior and child environment sub-domain scores (n=297).**

**Fig S2. Histogram of child environment sub-domain scores (possible score range: 0-60, n=297).**

